# MENTAL STATUS OVERVIEW IN THE ELDERLY DURING THE COVID-19 PANDEMIC: A PHYLOSOPHICAL PERSPECTIVE

**DOI:** 10.1101/2022.06.30.22277080

**Authors:** Andriani, Fransisca Lio, Ariyanti Saleh, Andi Muhammad Fiqri

## Abstract

**Background:** During the Covid-19 pandemic, there was a change in the mental status of the elderly, namely anxiety disorders and depression. It is important to know the mental status of the elderly and the role of effective communication because elderly people are easily infected with Covid-19. Therefore, it is necessary to carry out early detection in basic health services.

**Objective:** To know the description of the mental status of the elderly during the Covid-19 pandemic.

**Method:** Non-experimental quantitative design through a descriptive survey approach. The sample size is 202 people using quota sampling technique. Collecting data through home visits and implementing Covid-19 health protocols. The instruments used were the Geriatric Depression Scale 15 (Cronbach’s alpha 0.75) and the Geriatric Anxiety Scale (Cronbach’s alpha 0.93).

**Results:** There were 50.0% did not experience depression and 44.6% had mild depression. Although small, there is a small proportion of respondents experiencing moderate depression as much as 4.5% and 1.0% experiencing severe depression. There are 77.2% experiencing mild anxiety. A total of 21.8% experienced moderate anxiety. Although a small proportion, there are respondents experiencing severe anxiety as much as 1%.

**Conclusion:** During the Covid-19 pandemic, most of the elderly experienced a change in mental status.

**Suggestion:** Health workers are expected to carry out early detection of changes in mental status in the elderly.

## I. Background

Coronary Virus Disease 2019 (Covid-19) is a type of disease that was only recognized at the end of 2019 and has an immediate impact on the whole world. Virus type Severe Acute Respiratory Syndrome Corona Virus-2 (SARS CoV-2) originated from Wuhan City, Hubei Province, China (Cuartas-Arias, 2020). On March 11, 2020, World Health Organization (WHO) declared Covid-19 a pandemic. By June 2020, Covid-19 had spread to more than 200 countries, more than 6 million cases were reported with more than 370,000 deaths. When referring to the age group, the highest number of deaths in Indonesia is in the age group 60 years as much as 49.4% (Komite Penanganan Covid-19 dan Pemulihan Ekonomi Nasional, 2022).

Elderly people need special attention because at this time the individual experiences a decline in physical and psychological functions (Kementerian Kesehatan RI, 2019). Fear, worry, anxiety and feeling the threat of death in the elderly are increasing with news related to the increasing incidence of transmission and death due to Covid-19 (Keliat et al., 2020). The Covid-19 pandemic is a disaster that affects the mental status of individuals, including the elderly. It is estimated that approximately 80% of the total number of deaths due to Covid-19 are in people aged 65 years and there are around 37.1% experiencing anxiety disorders and depression during the Covid-19 pandemic (Lee et al., 2020).

In the current Covid-19 pandemic, early detection of the mental status of the elderly needs to be carried out as an early stage of preventing mental health and psychosocial problems in the elderly. Prevention of health problems needs to be done early as a form of early detection of worse things happening (Sherchan et al., 2017). There are 284 million people (3.76%) experiencing anxiety disorders and 264 million people (3.44%) experiencing depression, which is mostly experienced by elderly people (Ritchie H, 2017). In Indonesia, based on the results of the Basic Health Research (Riskesdas) it was found that the age group 65 years experienced depression as much as 8% and mental emotional disorders as much as 12.8% (Kementerian Kesehatan Republik Indonesia, 2018). The Covid-19 Quarantine House, Banyumas Regency, Central Java Province with 30 respondents found the results of 10 people (33.3%) experiencing mental emotional disorders, the most complaints were somatic complaints such as feeling anxious, tense (40%) and neglected daily activities (37%) (Nurjanah, 2020).

However, there have been no reports of the incidence of anxiety disorders and depression in the elderly during the Covid-19 pandemic. Based on the phenomenon that occurred, researchers felt that early detection was needed so they were interested in researching the mental status of the elderly during the Covid-19 pandemic.

## II. Material and Method

In this study, the study was approved by the Research Ethics Commission of Investment Integrated Services Office (registration ID: DPMPTSP.570/SKP.107/IV/2021). The design used is a non-experimental quantitative research through a descriptive survey approach. This type of design was chosen to be used in research because the researcher did not attempt to analyze and intervene on the variables studied but only provided an overview of the research objectives.

## III. Results

Respondents in this study were elderly people who managed to get health services during the Covid-19 pandemic from March 2020 to March 2021 and were then selected through the quota sampling technique with a total of 202 people. The total sample size in this study was 219 people and there were 17 people who refused to participate in the study including 4 people while sick, 13 people not willing to accept the researcher’s home visit. Research data was obtained through questionnaires filled out by respondents directly or with assistance and continued to carry out the Covid-19 pandemic health protocol which consisted of demographic data of respondents (age, gender, residential address, marital status, residence status, latest education, daily activities) and had received health services from the Public Health Centre, Geriatric Depression Scale 15, and Geriatric Anxiety Scale.

The Kolmogorov-Smirnov test and histogram were used so that the normality of the distribution of research data could be known with the result that the p value was <0.0001. From these results, it can be concluded that the distribution of the data in this study is not normal. Based on these results, the researchers present the results of this study in the form of frequency distribution tables and percentages based on the characteristics of respondents and research variables.

## IV. Discussion

### a. Phylosophy of depression in the elderly during the Covid-19 pandemic (Ontology, Epistemiology and Axiological Perspective)

In this study, the number of elderly people who experienced mild depression was significantly higher than the number of elderly people who experienced moderate or severe depression. The higher incidence of mild depression is possible because all respondents in this study had received health services from the puskesmas during the Covid-19 pandemic. These services are in the form of posyandu services for the elderly while still implementing the Covid-19 pandemic health protocol and direct medical visits by the elderly to the puskesmas.

The activity of the elderly in getting health services requires full support from the family so that it causes the elderly to meet each other. The meeting is an opportunity for the elderly to be able to tell each other while sharing experiences and sharing information related to Covid-19. In posyandu activities for the elderly, puskesmas officers provide various information related to the spread and prevention of Covid-19 transmission. Thus, elderly people can reduce the feelings of depression they feel during the Covid-19 pandemic.

In another study, it was found that the number of elderly people who experienced mild depression was more, namely 51 people compared to moderate depression, which was 5 people (Astuti, 2010). It was stated that this situation occurred because the participation rate of the elderly in participating in the posyandu for the elderly was very high due to good support from the family. By participating in posyandu activities, elderly people feel calm and will strengthen each other. The existence of good family support can make elderly people calm and have a good self-defense mechanism for solving any problems that arise. In another study related to family support, it was found that there was a significant relationship between family social support and the level of depression in the elderly (Parasari & Lestari, 2015). The incidence of depression in the elderly will be reduced if the family is able to provide high social support for the elderly.

During the Covid-19 pandemic, the posyandu for the elderly is an important activity to carry out as a means for the elderly to continue to receive health services with the help of posyandu cadres who have been provided with knowledge regarding the implementation of the posyandu for the elderly during the Covid-19 pandemic (Puspitaningsih et al., 2020). In another study related to posyandu for the elderly during the Covid-19 pandemic, it was found that health counseling activities related to preventing physical transmission can be carried out by puskesmas officers as an effort to reduce transmission of the Covid-19 virus and increase knowledge of the elderly about the Covid-19 pandemic (Herniwanti et al., 2020). Thus, it is hoped that the impact of the Covid-19 pandemic on changes in mental status can be minimized.

In this study, based on the marital status of most of the respondents, as many as 110 people were married, there were 59 married respondents who did not experience depression, 45 people experienced mild depression and two people experienced severe depression. Moderate depression is experienced by four people who are married and four people who are divorced. Moderate depression is experienced by four people who are married and 4 people who are divorced. There are 93 respondents who live with their families without depression, 85 people with mild depression, nine people with moderate depression and two people with severe depression. This illustrates that most of the respondents still have a life partner and family as a source of support. For the elderly who live alone, the source of support comes from the environment and the surrounding community. Elderly people who cannot adjust to changes that occur in their surroundings will be more at risk for experiencing changes in mental status.

More than half of the respondents in this study as many as 116 people feel unhappy for most of their lives. A small proportion of respondents as many as 12 people who feel dissatisfied with their lives. The emergence of feelings of being unhappy and dissatisfied with life in the elderly is due to the fact that elderly people feel that their freedom to carry out activities during the Covid-19 pandemic is too limited by their partners, especially by their families. Another thing that can also influence is the method used by the family in providing care to the elderly. Family members who live with elderly people spend more time outside the home to work and leave the elderly at home without giving freedom to activities. Even though they live at home with their family and are cared for and given attention by their children and grandchildren, they still feel lonely and sad so that depression in the elderly still occurs (Sari, 2020). The family as the closest person to the elderly can provide support in the form of attention, empathy, encouragement, giving advice, providing accurate information needed.

In this study, based on the latest education, the majority of respondents were elementary school graduates as many as 66 people. Respondents who graduated from elementary school as many as 33 people experienced mild depression and two people experienced severe depression. Moderate depression is experienced by respondents who did not finish elementary school and graduated from elementary school each as many as three people. During the Covid-19 pandemic, various news circulated from all news sources such as WhatsApp, Facebook, Twitter, YouTube, Tik Tok. The validity of the various information submitted is not necessarily reliable. Elderly people need to get accurate information related to everything related to the Covid-19 pandemic in order to avoid various unwanted things. The ability of elderly people to search for and interpret information varies depending on their education and knowledge. If the ability to find accurate information in elderly people is lacking, then the closest people such as spouses and family need to provide the information needed.

The last education taken by an elderly person can affect the incidence of depression (Pae, 2017). The level of education is closely related to the knowledge a person has. Education is a factor that can affect a person’s knowledge, because education can add insight so that actions and behavior based on knowledge will be better than those not based on knowledge. Elderly people who have low levels of education will have poor self-defense mechanisms in dealing with various problems in their lives.

In this study, there were 122 respondents who did not like staying at home and wanted to go out to do something new. Only a small number of respondents as many as 22 people feel that their lives are empty. There are 57 respondents who often feel bored. More than half of the respondents, i.e., 111 people, do not have a good spirit at all times. Although there are few, there are also 25 respondents who feel that the situation is hopeless. There were 37 respondents who were afraid that something bad would happen.

Various reports related to the Covid-19 pandemic have created feelings of fear of bad things that might happen in the lives of elderly people and can make elderly people think that there is no hope in life anymore. During the implementation of social interaction restrictions during the Covid-19 pandemic, the space for elderly people to move is limited. This limited space for movement can lead to feelings of boredom, life feels empty, and loss of enthusiasm. These various feelings that arise make elderly people feel uncomfortable staying at home and want to leave the house to do something new. Based on other research, there is a relationship between the Covid-19 pandemic and depressive disorders in the elderly in nursing homes (Rachmah & Tadjudin, 2021).

In this study, there were 24 respondents who felt they had a lot of problems with their memory when compared to other people. A small proportion of respondents as many as 47 people often feel helpless. Besides, as many as 20 people feel themselves useless. Almost half of the respondents, namely 70 people, think that many people are better off than themselves. Depression experienced can be caused because the elderly cannot adjust to the decline in all physical functions including decreased memory (cognitive) abilities. This decline in body functions can make elderly people feel that they cannot carry out daily activities such as when they were young, feel helpless and useless because they cannot carry out activities as before, which in turn can lead to feelings of inferiority and even isolation from the social environment. because they feel that there are many other people whose circumstances are better than the lives of the elderly.

Gymnastics exercises with light and moderate intensity can help improve cognitive function in the elderly (Listyasari, 2019). The physical condition of the elderly can still be maintained during the Covid-19 pandemic by carrying out elderly gymnastics which is held at the elderly posyandu and is very easy to carry out at home. Elderly gymnastics can make elderly people always feel fit. Physical exercise activities during the Covid-19 pandemic can be done at home using video as a medium (Aung et al., 2020). Physical exercise in the elderly can prevent the risk of falling and increase muscle strength and flexibility in the elderly. Elderly exercise can reduce the risk of various diseases such as hypertension, diabetes mellitus, coronary artery disease and the incidence of falls (Chang et al., 2020)

The incidence of depression in elderly people is more difficult to detect because elderly people often cover up feelings of loneliness and sadness by actively participating in activities such as recitation, social gathering, gymnastics and other activities outside the home (Livana et al., 2018). In this study, there were 160 respondents who continued to carry out many activities and interests or pleasures. Based on daily activities, there are 65 people who take care of the household who are not depressed and as many as 45 people have mild depression. Respondents who did not work as many as six people experienced moderate depression and as many as 2 people experienced severe depression.

During the Covid-19 pandemic, by continuing to carry out activities and interact with those around them, elderly people will be able to share stories and experiences and entertain each other. The relationship between the incidence of depression and social interaction is negative, meaning that the lower the level of depression, the better the level of social interaction (Kusumowardani & Aniek, 2014). Thus, interactions with others that are maintained among the elderly can reduce perceived depression.

### b. Phylosophy of anxiety disorders in the elderly during the Covid-19 pandemic (Ontology, Epistemiology and Axiological Perspective)

In this study, based on gender, it can be seen that 85 women experience more mild anxiety. This may be due to the fact that women are faster in finding and disseminating information related to Covid-19 in the community around the neighborhood where they live Therefore, women experience more anxiety in dealing with the Covid-19 pandemic than men, but this perceived anxiety can be reduced by providing health education regarding how to prevent the transmission of Covid-19 (Supriyadi & Setyorini, 2020).

In this study, more than half of the respondents, 112 people had difficulty sleeping and 119 people had difficulty starting to sleep. Difficulty getting to sleep and having trouble falling back asleep if you wake up in the middle of the night are indications of poor sleep quality. Poor sleep quality may occur because of worry, which is indicated by less than half of the respondents, as many as 92 people feel too worried about many things and 81 people cannot control the worries they feel. As many as 80 people felt that something terrible would happen to them and only 76 respondents had difficulty sitting still because they felt uncomfortable. In addition, as many as 110 people experienced abdominal pain, 109 people experienced muscle tension and 135 people experienced back pain, neck pain, or muscle cramps.

Various pain symptoms experienced by the elderly may be the cause of poor sleep quantity. Anxiety makes the mind confused, afraid, restless and uncomfortable, making it difficult for elderly people to start and or maintain sleep (Sohat et al., 2014). Action is needed so that feelings of anxiety and sleep problems experienced by the elderly are reduced or even resolved. During the Covid-19 pandemic there were symptoms of anxiety in the elderly including difficulty sleeping and difficulty starting to sleep (Arsy & Listyarini, 2021). Giving progressive muscle relaxation therapy in the elderly can help reduce muscle tension, improve sleep quality and can indirectly reduce body pain.

Based on the status of residence, there are 149 respondents who live with their families experiencing mild anxiety, 38 people experiencing moderate anxiety, and 2 people experiencing severe anxiety. In this study, there were 121 respondents who felt concerned about their children. Meanwhile, as many as 71 people are afraid of being a burden on their family or children. A total of 68 people who are less interested in doing something they usually enjoy. As many as 42 people feel they have no control over their lives.

During the Covid-19 pandemic, elderly people who live with their families (children and grandchildren) are given more attention by limiting the space for elderly people to move. The purpose of the family taking these restrictive measures is to protect the elderly because among family members, not all of them are free from Covid-19 disease. However, these restrictions can make elderly people feel they have no control over their own lives, feel unappreciated by their families, and feel like they are a burden to their families. All the feelings of the elderly can ultimately make the elderly become less interested in enjoying and carrying out their daily activities.

During the Covid-19 pandemic, families who live in the same house with elderly people generally feel anxious about various news about Covid-19 that is not yet known (Rayani & Purqoti, 2020). The family’s response to this feeling of anxiety is the emergence of excessive behavior towards the elderly, namely limiting social interaction with other family members who live in the same house.

In this study, there were 79 respondents who experienced a racing heart. 67 respondents experienced shortness of breath. as many as 84 people who feel like they have lost control or control. The feeling of fear of being judged by others was felt by 75 respondents. In addition, 57 respondents were afraid of being humiliated. Terdapat 110 orang yang mudah tersinggung. 100 respondents experienced anger easily. In addition, there were 84 people who felt easily startled or annoyed. Meanwhile, as many as 61 people feel separated or isolated from others. The difficulty to concentrate was experienced by 85 respondents.

The various complaints and symptoms experienced by the elderly require immediate treatment so as not to cause other unexpected things, such as depression and even suicide. Therefore, it is necessary to have a good understanding of the causes of feelings of anxiety, behavior of anxiety disorders, accurate information about Covid-19 and proper handling techniques are needed.

When feelings of fear, worry and anxiety are not clear, it can cause anxiety and can affect behavior changes, for example withdrawing from the environment, difficulty focusing on activities, difficulty eating, irritability, low ability to control emotions, sensitivity, inability to think using logic, and have trouble sleeping (Jarnawi, 2020). Psychosomatic symptoms that often appear can include skin allergies, shortness of breath, racing heart, cold sweats and nausea. Therapies that are easy to do to overcome anxiety are relaxation and Spiritual Emotional Freedom Technique (SEFT). SEFT therapy is a combination of Spiritual Power and Energy Psychology that can change the chemical conditions in the brain / neurotransmitters which can then change a person’s emotional state including depression. The results of this study indicate that relaxation and SEFT can help reduce various symptoms of anxiety disorders that are felt. In another study, it showed that there was an effect of SEFT therapy on reducing anxiety levels of narcotic users and addictive substance abuse (Dewi & Fauziah, 2018).

In this study, as many as 117 people feel concerned about their finances and 119 respondents feel concerned about their health. During the Covid-19 pandemic, elderly people can feel concerned about finances because of household economic conditions which may be disrupted due to restrictions on social interaction so that elderly people have to lose their jobs. Feelings of concern for health also arise because of the circulation of information related to Covid-19 which is more easily transmitted to elderly people with comorbidities and can even result in death. This needs to be considered by elderly family members because anxiety disorders can reduce body resistance and affect the co-morbidities experienced. The Covid-19 pandemic has an impact on physical health and mental health in particular, anxiety that can affect co-morbidities experienced by the elderly (Tobing & Wulandar, 2021). Families of elderly people are expected to be able to help the elderly to manage the anxiety they experience.

Based on the latest education, there are 52 respondents who did not finish elementary school who experienced mild anxiety and two people experienced severe anxiety. Respondents who graduated from elementary school as many as 18 people experienced moderate anxiety. The Covid-19 pandemic is a cause for concern. This feeling of anxiety can arise because the source of the information used is not necessarily clear. During the Covid-19 pandemic, everyone is able to receive information from various sources. The ability of elderly people to filter information related to Covid-19 varies depending on the education they have. One of the symptoms of anxiety is feeling worried that something bad will happen, feeling excessively worried, irritable, and difficult to relax (Setyaningrum & Yanuarita, 2020). The results of this study indicate that the Covid-19 pandemic has an influence on the mental status of the community, including causing excessive feelings of anxiety in individuals. One of the efforts to reduce this anxiety is to provide health education according to the needs of the community.

There is a difference in the level of anxiety about the transmission of the Covid-19 virus before and after being given education in the form of health education (Syamson et al., 2021). In addition, the results of other studies show that the anxiety felt by the elderly is the emergence of concerns related to physical health during the Covid-19 pandemic (Palgi et al., 2020). Therefore, this study shows that there are differences in the level of anxiety in elderly people who have been given an intervention in the form of health education and those who have not been given an intervention.

Based on daily activities, there are 93 people who take care of the household experiencing mild anxiety and as many as 19 people experiencing moderate anxiety. In this study, as many as 151 people felt tired. This feeling of tiredness arises because during the Covid-19 pandemic, elderly people who are used to working and doing activities outside the home end up having to stay at home more and this makes elderly people feel tired even though they are not active. Everyone can still maintain fitness, including the elderly through elderly gymnastics (Syahruddin, 2020). Thus, elderly people can carry out their daily tasks well without feeling excessive fatigue. Types of exercise that are suitable for elderly people include the stages of walking in place and going up and down a bench that is adjusted to the height (minimum 15 cm). This elderly exercise can improve the body’s defense system, reduce fatigue and create a sense of happiness in the elderly.

## V. Conclusion

The identification results showed that all respondents in this study experienced different occurrences of anxiety disorders including 156 people experiencing mild anxiety, 44 people experiencing moderate anxiety, two people experiencing severe anxiety and no respondents experiencing panic. In addition, during the Covid-19 pandemic, most of the elderly people also experienced changes in their mental status.

## Data Availability

All data produced in the present study are available upon reasonable request to the authors
All data produced in the present work are contained in the manuscript
All data produced are available online at

---

**Table 1.**
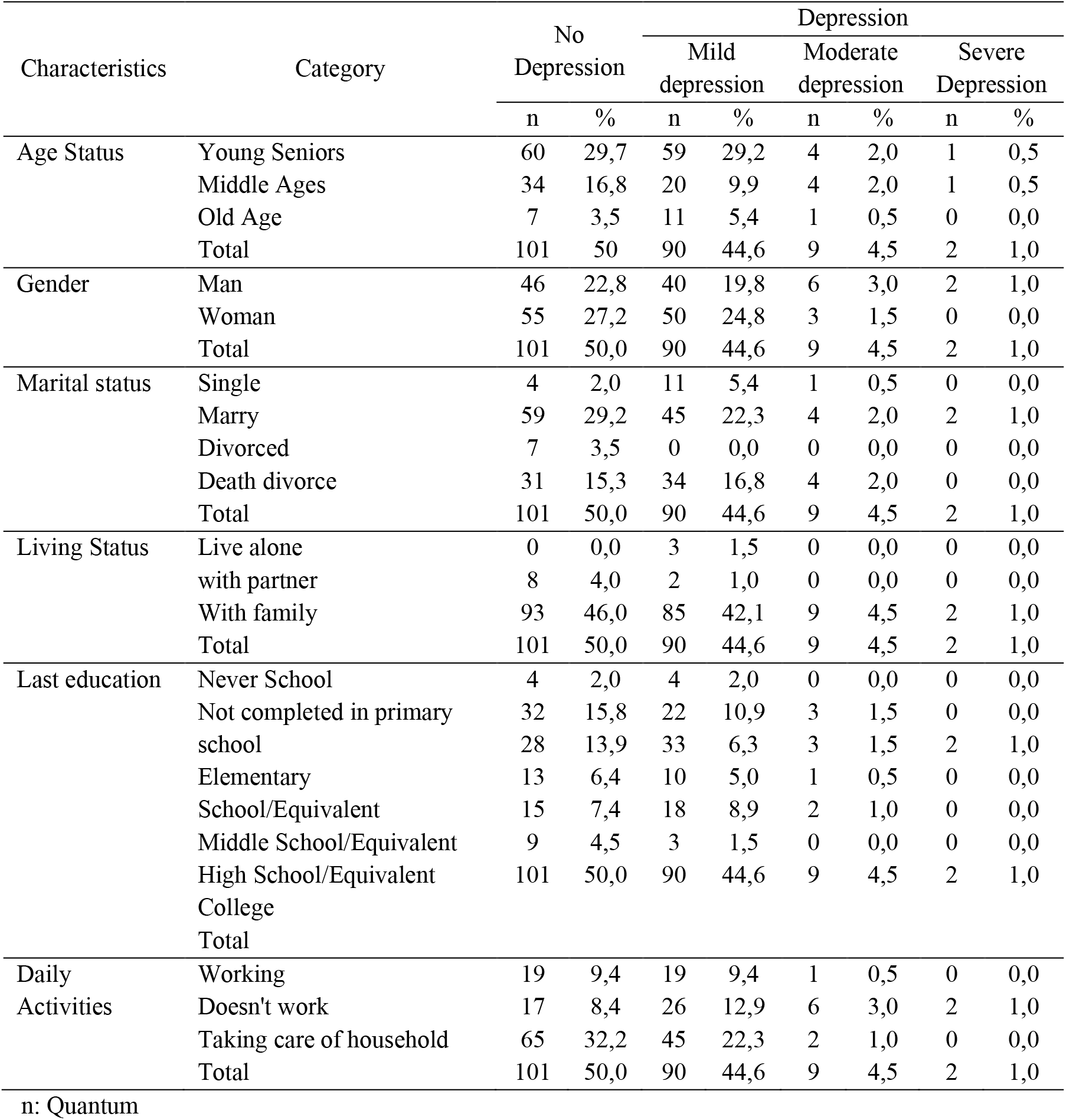
Distribution of Depression in the Elderly Based on Demographic Characteristics During the Covid-19 Pandemic

**Table 2.** The Incidence of Anxiety Disorders in the Elderly Based on Demographic Characteristics

| Characteristics | Category | Anxiety |  |  |  |  |  |
| --- | --- | --- | --- | --- | --- | --- | --- |
|  |  | Mild Anxiety |  | Moderate Anxiety |  | Severe Anxiety |  |
|  |  | n | % | n | % | n | % |
| Age Status | Young Seniors | 95 | 47,0 | 28 | 13,9 | 1 | 0,5 |
|  | Middle Ages | 47 | 23,3 | 12 | 5,9 | 0 | 0,0 |
|  | Old Age | 14 | 6,9 | 4 | 2,0 | 1 | 0,5 |
|  | Total | 156 | 77,2 | 44 | 21,8 | 2 | 1,0 |
| Gender | Man | 71 | 35,1 | 22 | 10,9 | 1 | 0,5 |
|  | Woman | 85 | 42,1 | 22 | 10,9 | 1 | 0,5 |
|  | Total | 156 | 77,2 | 44 | 21,8 | 2 | 1,0 |
| Marital status | Single | 14 | 6,9 | 2 | 1,0 | 0 | 0,0 |
|  | Marry | 85 | 42,1 | 25 | 12,4 | 0 | 0,0 |
|  | Divorced | 3 | 1,5 | 4 | 2,0 | 0 | 0,0 |
|  | Death divorce | 54 | 26,7 | 13 | 6,4 | 2 | 1,0 |
|  | Total | 156 | 77,2 | 44 | 21,8 | 2 | 1,0 |
| Living Status | Live alone | 0 | 0,0 | 3 | 1,5 | 0 | 0,0 |
|  | with partner | 7 | 3,5 | 3 | 1,5 | 0 | 0,0 |
|  | With family | 149 | 73,8 | 38 | 18,8 | 2 | 1,0 |
|  | Total | 156 | 77,2 | 44 | 21,8 | 2 | 1,0 |
| Last education | Never School | 2 | 1,0 | 6 | 3,0 | 0 | 0,0 |
|  | Not completed in primary school | 52 | 25,7 | 3 | 1,5 | 2 | 1,0 |
|  | Elementary School/Equivalent | 48 | 23,8 | 18 | 8,9 | 0 | 0,0 |
|  | Middle School/Equivalent | 17 | 8,4 | 7 | 3,5 | 0 | 0,0 |
|  | High School/Equivalent | 25 | 12,4 | 10 | 5,0 | 0 | 0,0 |
|  | College | 12 | 5,9 | 0 | 0,0 | 0 | 0,0 |
|  | Total | 156 | 77,2 | 44 | 21,8 | 2 | 1,0 |
| Daily Activities | Working | 31 | 15,3 | 8 | 4,0 | 0 | 0,0 |
|  | Doesn't work | 32 | 15,8 | 17 | 8,4 | 2 | 1,0 |
|  | Taking care of household | 93 | 46,0 | 19 | 9,4 | 0 | 0,0 |
|  | Total | 156 | 77,2 | 44 | 21,8 | 2 | 1,0 |

n: Quantum

